# Multimodal repertoire analysis unveils B cell biology in immune-mediated diseases

**DOI:** 10.1101/2022.01.04.22268769

**Authors:** Mineto Ota, Masahiro Nakano, Yasuo Nagafuchi, Satomi Kobayashi, Hiroaki Hatano, Ryochi Yoshida, Yuko Akutsu, Takahiro Itamiya, Nobuhiro Ban, Yumi Tsuchida, Hirofumi Shoda, Kazuhiko Yamamoto, Kazuyoshi Ishigaki, Tomohisa Okamura, Keishi Fujio

**Author notes:** Correspondence (M.O.), (K.F.).

## Abstract

**Objectives:** Despite the involvement of B cells in the pathogenesis of immune-mediated diseases, biological mechanisms underlying their function are scarcely understood. To overcome this gap, here we constructed and investigated a large-scale repertoire catalog of five B cell subsets of immune-mediated disease patients.

**Methods:** We mapped B cell receptor regions from RNA sequencing data of sorted B cell subsets. Our dataset consisted of 595 donors under immune-mediated diseases and health. We characterized the repertoire features from various aspects, including their association with immune cell transcriptomes and clinical features and their response to belimumab treatment.

**Results:** Heavy-chain complementarity-determining region 3 (CDR-H3) length among naïve B cells was shortened among autoimmune diseases. Strong negative correlation between interferon signature strength and CDR-H3 length was observed only in naïve B cells and suggested the role for interferon in pre-mature B cell development. VDJ gene usage was skewed especially in plasmablasts and unswitched-memory B cells of systemic lupus erythematosus (SLE) patients. We developed a scoring system for this skewing, and it positively correlated with peripheral helper T cell transcriptomic signatures and negatively correlated with the amount of somatic hyper mutations in plasmablasts, suggesting the association of extra-follicular pathway. Further, this skewing led to high usage of IGHV4-34 gene in unswitched-memory B cells, whose usage showed prominent positive correlation with disease activity in SLE. Gene usage skewing in unswitched-memory B cells was ameliorated after belimumab treatment.

**Conclusions:** Our multimodal repertoire analysis enabled us the system-level understanding of B cell abnormality in diseases.

## INTRODUCTION

B cells play critical roles in adaptive immunity, and their importance in immune-mediated disease (IMD) pathogenesis is suggested by the overlap of genetic risk variants of IMDs to regulatory regions in B cells ^1 2^ and by the clinical efficacy of B cell-targeted therapies in IMDs ^3 4^. Thus, deeper understanding of B cell biology could clarify IMD pathology.

To increase their diversity, B cell receptors (BCRs) are created by intrinsic genome-editing. In the bone marrow, one V, D and J gene for one clonotype is selected (VDJ recombination), and random deletions and insertions occur in the junctional regions. In the germinal center (GC), class switch recombination and loading of somatic hypermutation (SHM) further increases BCR diversity. These processes generate a vast variety of BCRs in an individual, a collection known as the “BCR repertoire”. Heavy-chain complementarity-determining region 3 (CDR-H3) is the most variable region and central to antigen specificity of BCRs.

IMDs consist of autoimmune and autoinflammatory diseases ^5^. Autoimmune diseases are characterized by two prominent features: the emergence of autoantibodies and hyperactivity of interferons (IFNs) ^5^, although the relationship between these features remains elusive. For creation of memory B cells and antibody secreting cells, affinity maturation through GC is a critical step. On the other hand, previous studies revealed that those cell types can be differentiated in a GC-independent manner ^6^, a process termed “extrafollicular” maturation. Pathogenic autoantibodies are generated through B cell extrafollicular reactions in systemic lupus erythematosus (SLE) and flares in SLE is characterized by the expansion of naive-derived activated effector B cells of extrafollicular phenotype ^7 8^. In addition, GC-independent B cell maturation was critical in monogenic lupus caused by a *TLR7* gain-of-function mutation ^9^, supporting the pathogenic role of extrafollicular pathway. The relevance of this B cell maturation process to disease pathology in various types of IMDs, as well as its association with intra-disease heterogeneity of SLE patients, could be better understood if placed in the context of a large-scale disease cohort. We recently created a gene expression database containing a large variety of immune cells from IMD patients as well as healthy volunteers ^2^. In this follow-up study, we focused on BCR features of 5 sorted B cell subsets that could be identified from RNA-seq data. We also examined their associations with transcriptomic and clinical information. Our multi-omics approach to a patient-derived, large-scale dataset generated new insights into B cell biology in immune-mediated diseases.

### Methods

Methods are provided in the supplementary information.

## RESULTS

### Profiling of BCR repertoires of IMD patients and healthy volunteers

We mapped BCR regions from RNA-seq reads of 5 sorted CD19^+^ B cell subsets: CD27^−^IgD^+^ naïve B cells, CD27^+^IgD^+^ unswitched memory B cells (USM B), CD27^+^IgD^−^ switched memory B cells (SM B), CD27^−^IgD^−^ double negative B cells (DN B) and IgD^−^CD27^++^CD38^+^ plasmablasts (**Fig. 1A**). We extended our cohort from the previous report ^2^ and samples as follows: 136 patients with SLE, 90 patients with systemic sclerosis (SSc), 85 patients with idiopathic inflammatory myopathy (IIM), 147 patients with other IMDs and 137 healthy controls (HC), giving a total of 595 participants (**Fig. 1A**). For 22 SLE patients, samples were also obtained after a half-year treatment with belimumab, a biologic targeting B-cell activating factor (BAFF). After mapping and quality control, 5.8 million productive unique CDR-H3 clonotypes were identified from 2,893 samples. On average, 2,801 distinct productive clonotypes were identified in each sample (**Fig. S1A**). More than 5,000 BCR-derived reads were identified per million RNA-seq reads in each subset (median), and in plasmablasts it reached 1.1 × 10^*5*^ reads (**Fig. 1B**), reflecting their specialized role in secreting antibodies.

**Fig. 1.**
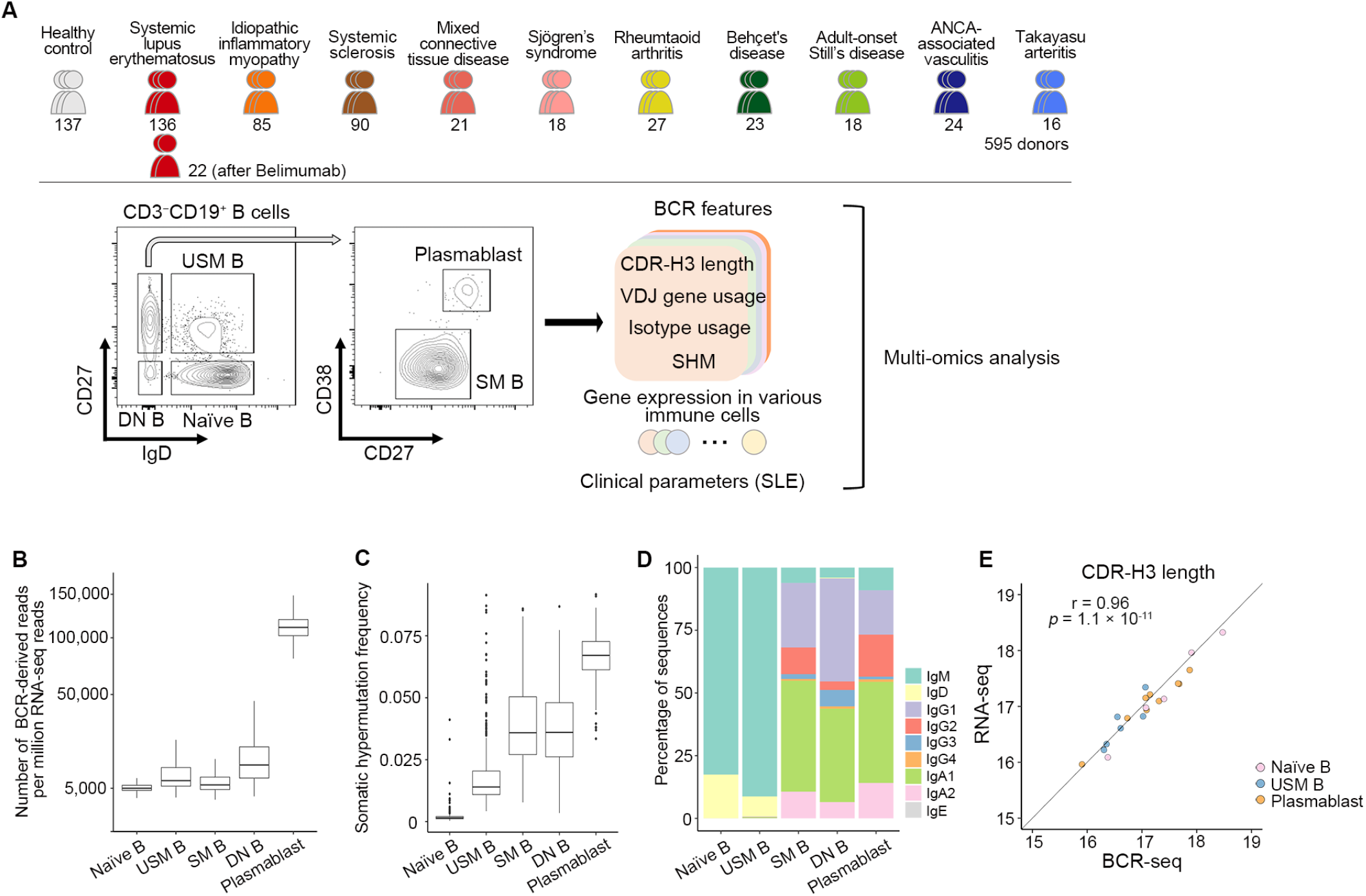
Study design and quality features of our dataset. **(A)** Study design used in this report. Sample numbers after quality control are presented under each disease. **(B)** Number of RNA-seq reads aligned to BCR regions per million reads in 5 B cell subsets. **(C)** Comparison of the per base mutation frequency in the V gene region of 5 B cell subsets. **(D)** Comparison of median isotype frequencies in each subset among healthy controls. **(E)** Comparison of CDR-H3 region length between the results from RNA-seq experiment and BCR-seq experiment. Solid line indicates y = x.

As naïve B cells are expected to have few SHM, the mutation frequency can reflect the error rate in PCR or sequencing steps. The SHM frequency in V genes in naïve B cells was well controlled at 0.15% (median), in clear contrast to 6.7% (median) in plasmablasts (**Fig. 1C**). Equivalent to the previous single cell BCR analysis that classified more than 99% of IgG+ and 98% of IgA+ cells as memory cells ^10^, IgG or IgA were rarely detected (0.4%) in naïve B cells in our dataset (**Fig. 1D**). Together, our dataset captured biologically reasonable characteristics of BCR repertoires.

In order to evaluate the influence of the number of reads for BCR repertoire characterization, we performed targeted BCR sequencing (BCR-seq) for 20 samples which also have been undergone RNA-seq. Despite the difference in read depth (Fig. S1B) and the number of identified CDR-H3 clonotypes (Fig. S1C), average CDR-H3 length (Pearson r = 0.96, Fig. 1E) and VDJ gene usage (r = 0.97, Fig. S1D) were very close between RNA-seq and BCR-seq. These results indicated that bulk RNA-seq of sorted B cells have enough information to estimate these relative features while not capturing whole clonotypes. Thus, in our following RNA-seq based BCR repertoire analysis, we focused on the relative features rather than whole clonotypes.

### In autoimmune diseases, CDR-H3 length is shortened in naïve B cells in an interferon signal strength-dependent manner

CDR-H3 loop length is one of the classic indices for the assessment of repertoire bias ^11^. When we compared CDR-H3 length with HC, marked shortening of CDR-H3 lengths in naïve B cells (by more than 1 amino acid) and DN B cells among autoimmune diseases were observed (**Fig. 2A**; **Fig. S2A**). Conversely, CDR-H3 lengths in USM B cells and plasmablasts of SLE patients were longer than those in HC (**Fig. 2A**; **Fig. S2A**). Among the sections of the CDR-H3 region, lengthening in plasmablasts was observed in germline-coded regions but not in junction regions, in contrast to shortening in overall sections in naïve B cells (**Fig. 2B**; **Fig. S2B**).

**Fig. 2.**
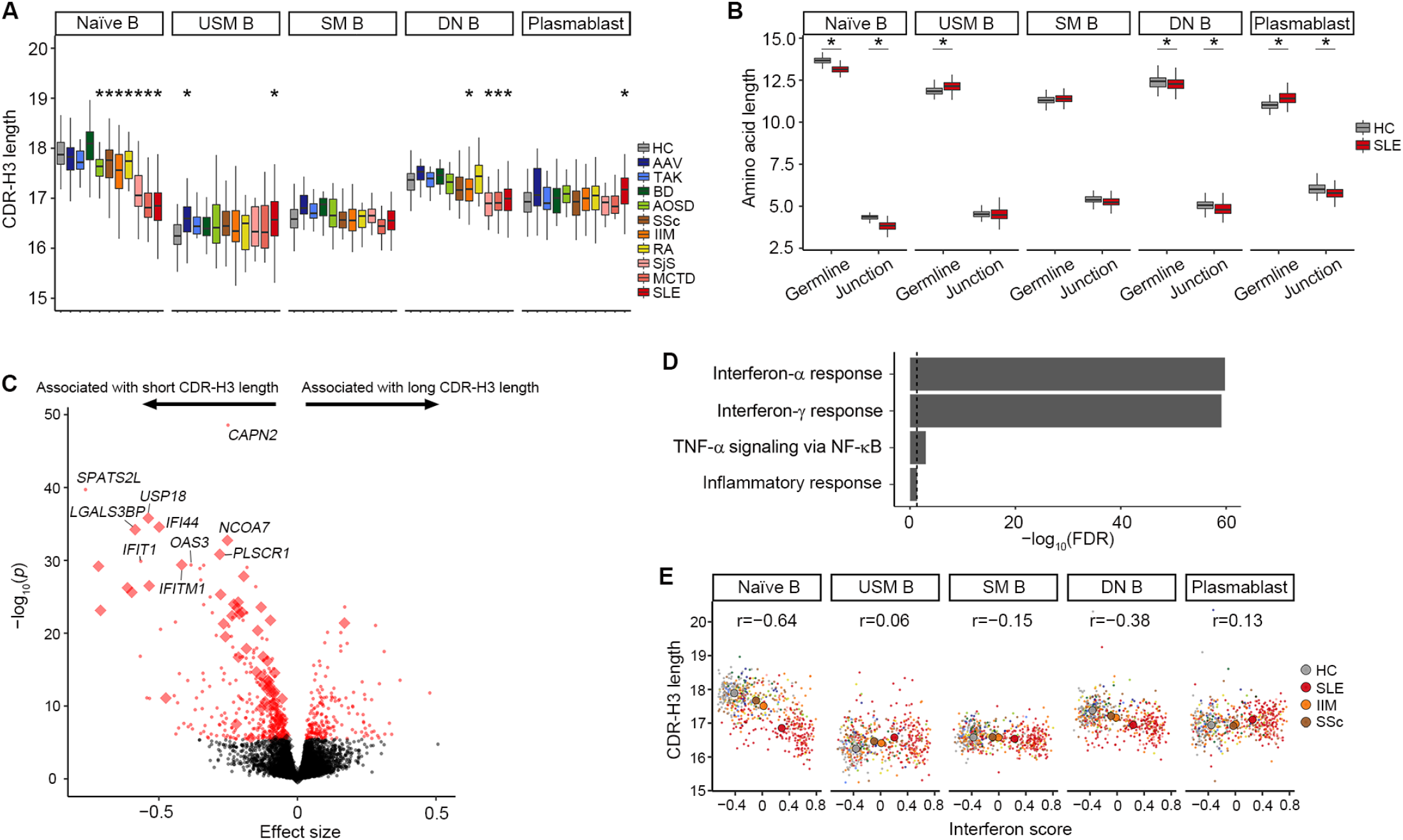
In autoimmune diseases, CDR-H3 length is shortened in naïve B cells in an interferon signal strength-dependent manner. **(A)** Comparison of CDR-H3 length between IMDs and HC. The difference between HC and each disease was tested with linear regression, adjusting for age and batch effects. *, *p* < 0.05/50 compared to HC. **(B)** Comparison of germline and junction part length of CDR-H3 region (see also Fig. S2B). The differences between HC and SLE were tested with linear regression, adjusting for age and batch effects. *, *p* < 0.05/10. **(C)** Association of naïve B cell gene expression with naïve B cell CDR-H3 length. Fixed effect meta-analysis *p* values and effect sizes are plotted. Genes that attained Bonferroni-adjusted *p* < 0.05 are marked red. Genes in “HALLMARK_INTERFERON_ALPHA_RESPONSE” gene set from MSigDB are marked with diamonds. **(D)** Gene set enrichment analysis. Results of genes significantly negatively associated with CDR-H3 length in naïve B cells. FDR, false discovery rate. **(E)** Correlation between IFN score and CDR-H3 length in 5 B cell subsets. Mean values among HC, SLE, SSc, and IIM are marked with large points. The color of each point represents the clinical diagnosis as illustrated in Fig. 1A. Pearson’s r is provided.

Observations so far were only from productive sequences. Non-productive sequences, which refer to the sequences with a frameshift or stop codon, can be used as a means of studying the pre-selection repertoire ^12^. In our dataset, shortening of CDR-H3 lengths among autoimmune diseases was observed in non-productive sequences of naïve and memory B cells (**Fig. S2C**). Thus, together with the observed shortening of overall CDR-H3 sections in productive sequences, short CDR-H3 clonotypes may be generated before selection of mature naïve B cells (e.g., during the process of VDJ recombination and nucleotide insertion/deletion at the junctions). This observation is analogous to previous reports of shortening of CDR3 length in pre-selection T cell receptors among type 1 diabetes patients ^12^. Moreover, CDR-H3 lengthening in germline regions of plasmablasts in SLE may be the consequence of skewed VDJ gene usage as described in the next section.

CDR-H3 shortening was especially prominent in naïve B cells from diseases with high IFN signatures (**Fig. 2A**; **Fig. S2D**), although CDR3 length was not as divergent among light chains (**Fig. S2E**). To investigate further, we tested the association of gene expression and CDR-H3 length in naïve B cells. Here, in order to avoid the confounding of disease effects (Simpson’s paradox), we tested the association in each disease and subsequently performed meta-analysis (see **Methods**). Genes associated with a short CDR-H3 length were significantly enriched in IFN-stimulated genes (ISG, **Fig. 2C, D, Table S1**), and the association was observed in each autoimmune disease (**Fig. S2F**). CDR-H3 length in naïve B cells was also significantly associated with ISG expression in 4 other B cell subsets, indicating the association of generalized IFN activity itself with CDR-H3 length (**Fig. S2G, Table S1**). Strong negative correlation between CDR-H3 length and IFN activity was characteristic of naïve B cells among 5 B cell subsets (**Fig. 2E**). These results suggested a role of IFN signaling at the early stage of B cell development. This is consistent with previous findings of IFN-induced abnormal B cell maturation in bone marrow of SLE ^13^ and further suggested its relevance in a wide range of autoimmune diseases.

### Skewed gene usage in autoimmune diseases suggested an abnormal repertoire maturation process

It was previously suggested that there was differential usage of VDJ genes according to B cell subsets in humans and mice ^14 15^, but the generalizability of this finding in a large population has been elusive. In our cohort, gene usage preference among 5 B cell subsets was also observed (**Fig. S3A**). More surprisingly, by principal component analysis (PCA) of VDJ gene usage, samples were clearly separated according to cell types (**Fig. 3A**). This observation is analogous to previously reported clear separation of 3 B cell subsets by PCA of VDJ use in C57BL/6 mice ^15^. Those results suggested that B cell selection was based on VDJ gene segments independent of the sequence of their antigen-binding regions in humans.

**Fig. 3.**
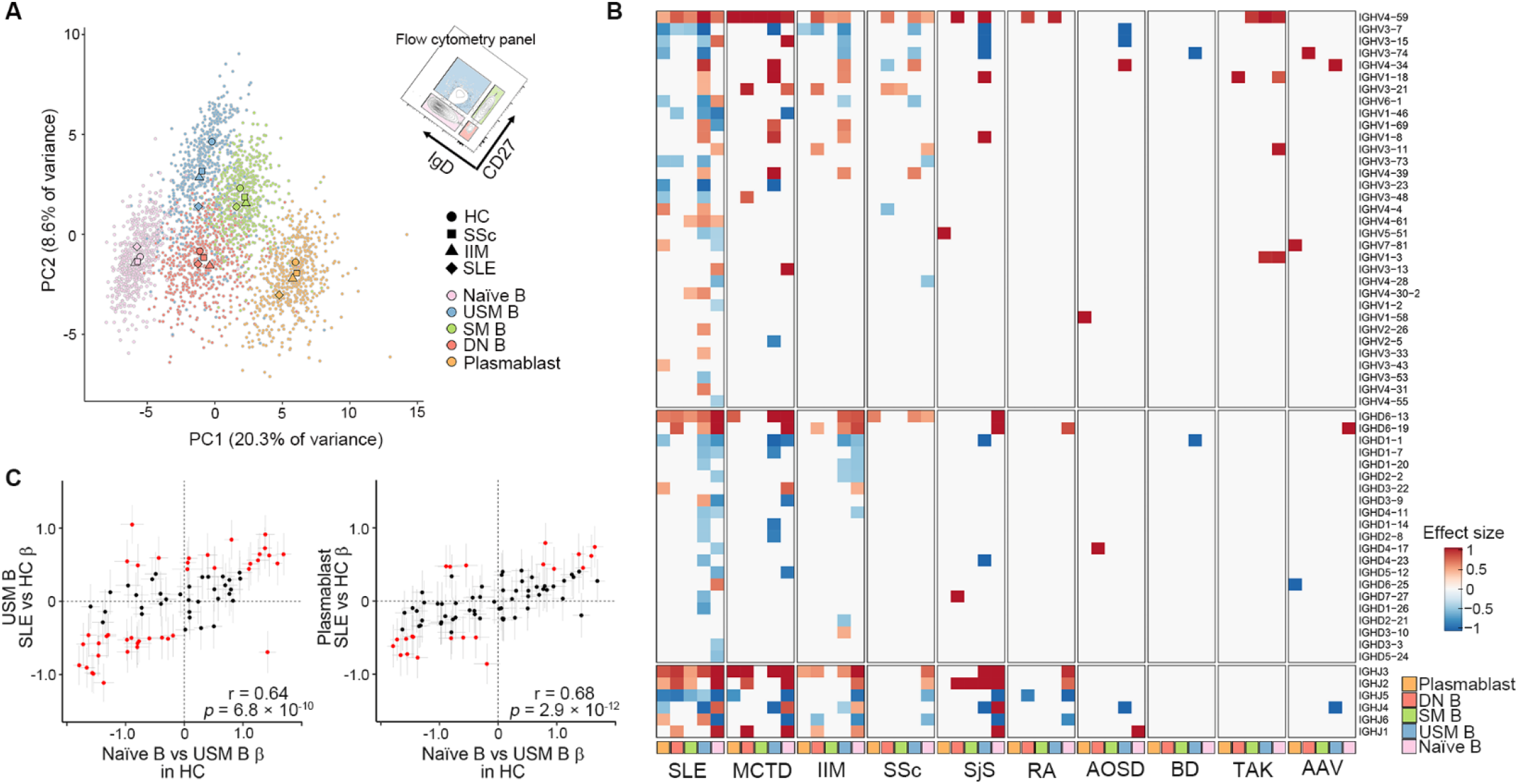
Skewed gene usage in autoimmune diseases suggested an abnormal repertoire maturation process. **(A)** Principal component analysis plot of all samples based on VDJ gene usage. Centroids of HC, SSc, IIM, and SLE in each subset are marked with large points. **(B)** Skewed gene usage in IMDs compared to HC. In each disease-subset (column), gene usage (row) was compared with HC after adjusting for age and batch effects. Effect sizes of linear model comparison are plotted if the difference was statistically significant (FDR < 0.05 after Benjamin-Hochberg correction). Only genes that attained FDR < 0.05 in at least one comparison are shown. **(C)** Comparison of β estimates in usage comparison between naïve B vs USM B in HC (x axis) and SLE vs HC (y axis) in USM B (left) or plasmablasts (right). Each plot represents each VDJ gene. Genes that attained FDR < 0.05 in SLE vs HC comparison are marked red. Grey bars indicate 95%CI. Pearson’s r and its significance are provided.

It has been shown that there is skewed usage of the IGHV genes in IMDs ^16^ with dependence on the stage of disease ^17^, although the sample size has been limited. In case-control comparisons in our dataset, many differentially used genes were identified especially in autoimmune diseases rather than autoinflammatory diseases (**Fig. 3B, Table S2**). Notably, in the PCA space the gene usage signature was drastically skewed into the 7 o’clock direction in USM B and plasmablasts of SLE patients. (**Fig. 3A**; **Fig. S3B**). This direction was in parallel with the vector between USM B and Naïve B subsets among HC. Indeed, VDJ gene usage differences between SLE and HC in USM B, as well as plasmablasts, showed clear agreement with gene usage differences between naïve B and USM B in HC (**Fig. 3C**). Regarding IIM and SSc, the repertoire skewed in the same direction in the PCA space in USM B, but not obviously in plasmablasts (**Fig. 3A; Fig. S3B**). Motivated by these observations, we hypothesized that gene usage difference between B cell subsets among healthy controls can be utilized as functional references in BCR repertoire analysis, analogous to “pathways” widely used for transcriptome analysis. We developed a score to quantify such skewing based on VDJ gene usage patterns of B cell subsets. The score is calculated as the weighted sum of normalized VDJ gene usage of each sample, and the weights were defined by the usage difference between two subsets in HC (see **Methods, Table S3**). We calculated 10 patterns of such scores for each sample with possible combinations of 5 B cell subsets. Among them, the score calculated based on naïve B versus USM B comparison most clearly separated SLE patients from HC (AUC=0.933, **Fig. S3C**), although the scores based on DN B versus SM B or DN B versus USM B comparisons showed equivalent performance (AUC=0.920, 0.931). Based on these observations, we considered that the gene usage transition from CD27 negative (i.e., naïve B and DN B) to positive (i.e., USM B, SM B and plasmablasts) populations may be distorted in CD27 positive populations in SLE and other IMDs. Thus, hereafter we refer to the score calculated based on naïve B versus USM B gene usage difference in HC as reference as “Repertoire Naïveness (RN) score” and utilize it as a metric of repertoire abnormality in IMDs.

Consistent with the appearance in PCA, our RN score was the highest among SLE patients in USM B or plasmablasts (**Fig. S3D**). To validate our approach, we adopted the weights and scoring system from our dataset to the BCR-seq data derived from sorted B cells from human tonsils in a previous study ^18^. The RN score was clearly high in naïve B cells and low in memory B cells in this independent dataset as well as in our dataset (**Fig. S3E, F**).

As reported previously ^19^, the use of each VDJ gene was associated with different CDR-H3 lengths (**Fig. S3G**). The genes associated with an extended CDR-H3 in plasmablasts among HC were preferentially used by plasmablasts in SLE (**Fig. S3H**). CDR-H3 extension in SLE was observed only in regions coded by VDJ genes (**Fig. S2B**). Thus, the extended CDR-H3 length in SLE in this subset is seemingly associated with abnormal VDJ gene usage.

### The repertoire abnormality in autoimmune diseases is associated with the extrafollicular pathway

Extrafollicular maturation of plasmablasts has been reported as a hallmark of SLE ^7^. Next, we assessed the association of the observed “naïve like” repertoire and the extrafollicular maturation. Although extrafollicular maturation-derived antibody secreting cells undergo SHMs, they are expected to have less SHMs ^6 8 20^ compared to GC-derived cells. In line with this notion, the frequency of SHMs in plasmablasts was significantly lower in SLE patients than in HC (**Fig. S4A**). Furthermore, it showed a significant negative correlation with the RN score (**Fig. 4A**). To examine the relationship in clonotype levels, we divided BCR clonotypes into bins according to the frequencies of SHMs and calculated RN scores in each bin. In this comparison, we observed that clonotypes with a lower number of mutations showed higher RN scores (**Fig. 4B**). Taken together, naïve-like gene usage in plasmablasts is associated with a low load of SHM. Reflecting the accumulation of mutations with aging ^21^, SHM frequencies in plasmablasts showed a significant positive correlation with aging (*p* < 2 × 10^−16^), although the RN score was not affected by aging (*p* > 0.05, **Fig. S4B**). The RN score distinguished SLE patients from HC more accurately than did the SHM frequency (**Fig. S4C**). Thus, although well correlated, the RN score might reflect disease-associated pathways better than the SHM frequency.

**Fig. 4.**
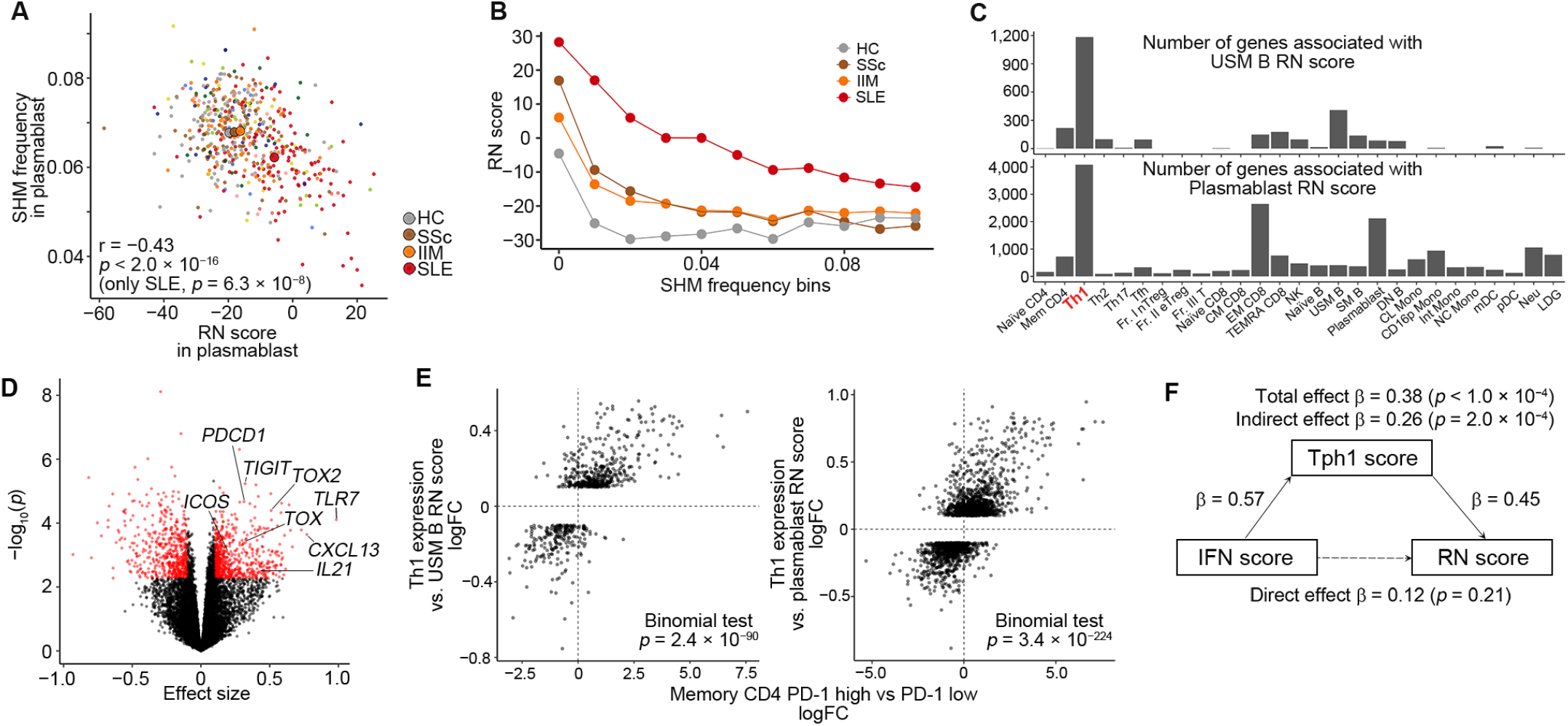
The repertoire abnormality in autoimmune diseases is associated with the extrafollicular pathway. **(A)** Correlation of RN score and SHM frequency in the V gene region of plasmablasts. *P* values were calculated with linear regression adjusting for diagnosis, age and batch effects. Mean values among HC, SLE, SSc, and IIM are marked with large points. The color of each point represents the clinical diagnosis as illustrated in Fig. 1A. **(B)** RN scores of plasmablast clonotypes stratified by SHM frequency. In each disease, clonotypes for which ≥ 200 nucleotides in the V gene were sequenced were aggregated and divided into bins according to mutation frequency in the V gene region. In each bin, RN score was calculated based on VDJ gene usage. **(C)** Number of significantly associated genes with RN score in USM B (top) and plasmablasts (bottom) among expressed genes in each immune cell among SLE patients. Full description of subset names is in Table S5. **(D)** Association of gene expression in Th1 cells and RN score among SLE patients. RN score was calculated in USM B. Significantly associated genes are colored red. **(E)** Comparison of the effect size estimates of Th1 expression versus RN score comparison in our cohort (Y-axis) and PD-1^hi^ versus PD-1^**−**^ T cells comparison in the data by Rao, *et al* ^22^ (X-axis). Only genes significantly associated with RN scores in our cohort were included for the analysis. *P* values were calculated by the binomial test for concordance of the direction of effect size estimates. logFC, log fold change. **(F)** Mediation model representing the relationships among IFN score, Tph1 score and RN score in plasmablasts among SLE patients. The bootstrapped indirect effect was 0.26 (95% CI, 0.14-0.39; *p* = 2 × 10^**−**4^). Thus, the indirect effect was statistically significant.

We next compared the RN scores of 136 SLE patients focusing on the gene expression of 27 immune cell subsets that were obtained from the same patients. The number of genes that showed a significant correlation with the RN score was the largest in Th1 cells for both plasmablasts and USM B (**Fig. 4C**). The top associated genes in Th1 cells included *PDCD1, TOX* and *ICOS*, which are hallmarks of peripheral helper T cells (Tph) ^22^ (**Fig. 4D, Table S4**). Indeed, the Th1 genes associated with RN score showed significant concordance with the direction of change in the differential expression analysis between PD-1^hi^ versus PD-1^−^ T cells from the original report ^22^ (**Fig. 4E**). Notably, association of Tph gene expression with RN score was validated in an independent SLE cohort^23^, suggesting the robustness of this association and high quantitative performance of RN score **(Fig. S4D, E)**. Thus, the Th1 genes associated with the RN score may reflect the expansion of Tph-like subpopulation among Th1 cells. The Tph gene expression score in Th1 cells (Tph1 score) was high in SLE (**Fig. S4F**), consistent with the reported expansion of Tph population ^24^, particularly with Th1 phenotype ^25^, in SLE. The Tph1 score showed a strong correlation with the RN score among SLE patients, both in USM B and plasmablasts (**Fig. S4G**). Correlation of these two scores was also prominent in USM B in IIM, although the correlation in plasmablasts was lower than that in SLE. These results may reflect the shared and distinct pathogenicity among autoimmune diseases. The association of Tph1 score and plasmablast RN score was also validated in our independent SLE cohort ^23^ (**Fig. S4H**). Although IFN signal strength was also correlated with RN scores (**Fig. S4I**), this association was mostly mediated by the Tph1 score in the mediation analysis (**Fig. 4F**). The direct effect of IFN signal strength on RN score was not significant (*p* = 0.21) after considering indirect effect via Tph1 score (**Fig. 4F**). The proposed function of Tph cells is that they provide help to B cells outside the GC ^22 24^. Collectively, associations of RN score with low SHM frequencies and high Tph1 score support the notion that the naïve-like repertoire in IMDs, especially in SLE, is a consequence of excess extrafollicular maturation.

In addition, the RN score of USM B correlated significantly and positively with the expression of *TBX21* in USM B, which codes for T-bet (**Fig. S4J**). As T-bet^**+**^ B cells reportedly have distinct functions ^26^ and are associated with autoimmunity ^27^, this naïvenesss-associated USM B population might play a role in autoimmune pathogenesis.

### Repertoire abnormalities are associated with disease activity in SLE

We next tested the association of BCR repertoire abnormalities and disease activity in SLE. RN scores in plasmablasts and USM B showed significant positive correlations with SLEDAI-2K, a measure of SLE disease activity (**Fig. 5A**), and these associations were still significant after adjusting for Tph1 scores (*p* = 1.3 × 10^−3^, 1.7 × 10^−3^) and ISG scores (*p* = 5.3 × 10^−4^, 2.3 × 10^−5^), indicating the clinical relevance of RN scores. In addition, RN score in plasmablasts significantly correlated with total IgG concentrations in sera (**Fig. S5A**).

**Fig. 5.**
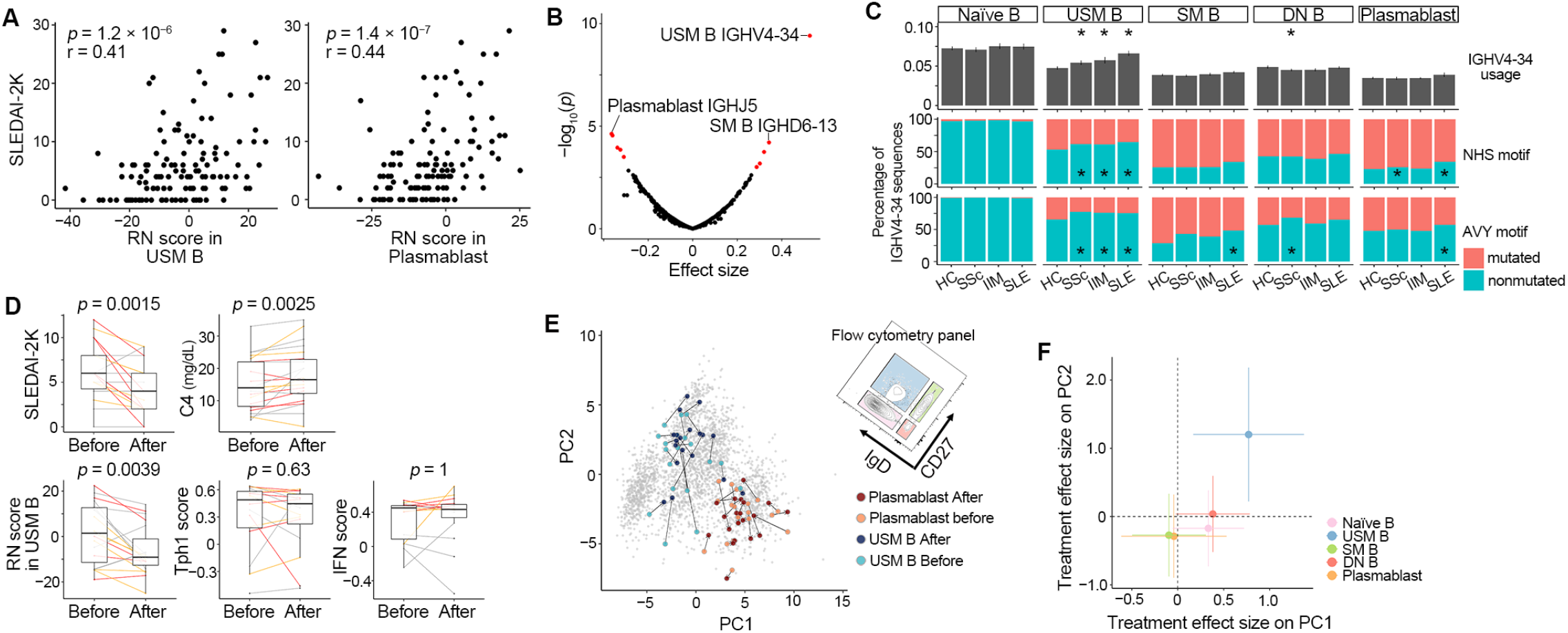
Repertoire abnormalities are associated with disease activity in SLE and attenuated by belimumab treatment. **(A)** Correlation between RN scores and SLEDAI-2K among SLE patients. Pearson’s r and its significance are provided. **(B)** Association of VDJ gene usages among 5 B cell subsets and SLEDAI-2K. Significantly associated genes (FDR < 0.05) are red. **(C)** Comparison of mean IGHV4-34 usage (top) and percentage of clonotypes with mutations in the NHS motif (middle) or the AVY motif (bottom) among IGHV4-34 clonotypes. For usage comparison, the differences between HC and each disease were tested with linear regression adjusting for age and batch effects. Bars indicate 2SE. For mutation comparison, Fisher’s exact test comparing the number of mutated/nonmutated clonotypes between HC and each disease was performed in each subset. *, *p* < 0.05/15. **(D)** Comparison of clinical parameters and scores before and after treatment with belimumab. Patients are colored according to improvement in SLEDAI-2K after treatment (red, ≥ 4; orange, 1-3; grey, ≤ 0). Paired Wilcoxon signed-rank test. **(E)** Change in BCR repertoire after belimumab treatment. Paired samples before and after treatment were connected on the same PCA space as Fig. 3A. **(F)** Belimumab treatment effects on PC scores by fitting the linear mixed model. Bars indicate 95% CI.

We next asked whether specific V, D or J gene usage was associated with disease activity. In SLE, among the 5 B cell subsets, IGHV4-34 usage in USM B showed by far the most significant positive association with SLEDAI-2K (*p* = 3.9 × 10^−10^, **Fig. 5B, Fig. S5B**). Although the expansion of antibody secreting cells with IGHV4-34 in acute SLE has been reported ^8^, the clinical relevance of its usage in USM B has not been clarified. IGHV4-34 is a V gene that is predominantly used in naïve B cells (**Fig. S3A**) and its usage explained 28% of the variance of RN score in USM B (**Fig. S5C**). Mediation analysis indicated that over one-half of the association between RN scores in USM B and SLEDAI-2K was mediated by IGHV4-34 usage. This contrasted with the independent direct association of RN scores in plasmablasts with SLEDAI-2K (**Fig. S5D**). Thus, repertoire naïveness in SLE results in high usage of IGHV4-34 in USM B and that seems to be associated with disease activity.

It has been reported that AVY and NHS motifs in IGHV4-34-coded amino acid sequence recognize autoantigens and are associated with autoimmunity ^28^. Mutations in these motifs abrogate its autoreactivity and only such clones without autoreactivity are selected for maturation under healthy conditions ^28 29^. In our dataset, AVY and NHS motifs were significantly more conserved without mutations in SLE USM B compared to HC as well as in plasmablasts (**Fig. 5C**). Thus, IGHV4-34 clonotypes in SLE might feature binding to autoantigens.

### Belimumab attenuated abnormal repertoire in SLE

For SLE, the BAFF inhibitor belimumab is one of the two biologics that has been approved by the United States Food and Drug Administration ^3^. In SLE, BAFF is induced by type ? IFN stimulation and supports the survival and differentiation of autoreactive B cells ^30 31^. To assess the effect of BCR modifications, we tested the BCR repertoire before and after treatment with belimumab in 22 cases of SLE. Belimumab effectively reduced SLEDAI-2K and increased complement levels in our cohort (**Fig. 5D**). In order to assess the overall trend, we projected cases before and after treatment on the same PCA space as Fig. 3A. Belimumab treatment significantly changed the repertoire of USM B against the naiveness-associated direction, but not in the other subsets (**Fig. 5E, F**). Subsequently, belimumab significantly reduced the RN scores in USM B (*p* = 0.0039), although we detected no effects on Tph1 scores or IFN scores (**Fig. 5D**). Thus, belimumab had the most obvious effect on repertoire naïveness in USM B. After treatment, we observed the trend of reduced fraction of IGHV4-34 in USM B and significantly reduced fraction of nonmutated AVY/NHS motifs in USM B and plasmablasts (**Fig. S5E**). These changes in the autoimmune-associated fraction of BCRs are supposed to be the consequence of the amelioration of repertoire naïveness in USM B.

## DISCUSSION

In this report, we characterized BCR repertoire abnormalities in IMDs using a multi-layer dataset.

We identified marked shortening of CDR-H3 length in naïve B cells of autoimmune disease patients in an IFN signal strength-dependent manner, as well as lengthening in plasmablasts of SLE patients. Shortening of BCR CDR-H3 length in peripheral blood mononuclear cells of SLE patients had been reported ^32^. In contrast, antibodies with long CDR-H3 length were associated with autoreactivity ^33^ and long CDR-H3 length in class switched clonotypes in SLE was also reported ^16^. Our results are consistent with all of these seemingly contradicting reports and reinforce the importance of using sorted B cells for repertoire analysis. In healthy conditions, CDR-H3 length are shorter in memory than naïve subsets both in T and B cells ^34 35^ and long CDR-H3 clones might be under negative selection pressure ^35^. In our study, short CDR-H3 length in naïve B cells among autoimmune disease patients resulted in small length gap between naïve and memory B cells compared to HC (**Fig. 2A**), which might mitigate the repertoire selection pressure, leading to the persistence of autoreactive clones in memory B cells.

In gene usage analysis, we observed a global pattern of skewness in specific subsets of IMDs as quantified with the RN score. This score can measure extrafollicular maturation based on the observation of its association with naïve-like gene usage, low SHM load and its correlation with Tph signature gene expression in Th1 cells. The observed significant association of these scores with disease activity and their responses to clinically effective treatment suggested the clinical relevance of the gene usage skewness we observed in this study. Although the involvement of extrafollicular maturation in acute SLE patients has been reported^8^, its relevance to a wide range of IMDs, as well as intra-disease dynamics among SLE patients, have not been clearly understood. Our development of gene usage-based scoring system for extrafollicular maturation activity enabled us to quantify and compare it in a large cohort. Interestingly, in SLE patients, the repertoires of USM B and plasmablasts seemed to be skewed similarly in response to Tph expansion. This was in contrast to the observation of similar associations in USM B but less so in plasmablasts in IIM. This result may suggest the existence of a “gatekeeper” for plasmablast maturation that is defective in SLE. Together with CDR-H3 length analysis, our results support the notion that the break of tolerance occurs both in central and peripheral checkpoints in SLE ^36 37^. Also, our analysis supported direct and indirect associations of IFN signaling with both of these checkpoints.

We observed that belimumab treatment decreased RN score in USM B. Previous study demonstrated that patients treated with belimumab more than 7 years had the reduced fraction of IGHV4-34 among unmutated IGM clonotypes compared to controls ^38^. Our result is largely consistent with this finding, but we observed more obvious effects of belimumab on RN score than on IGHV4-34 fraction in USM B after half-year treatment. This result may suggest the effect of belimumab on extrafollicular pathway of USM B, rather than specific autoimmune clonotypes. As repertoire in USM B was similarly skewed in other autoimmune diseases as well, it could have potential treatment effects on these diseases, although further investigation of the pathogenicity of this pathway is required.

We need to acknowledge some limitations in our study. First, although our RNA-seq based approach has an advantage of studying large sample size, it is limited in its sequencing depth compared to BCR-targeted sequencing. Although RNA-seq of sorted B cell subsets had enough information to assess the relative repertoire features, for deep characterization of clonotype level abnormalities such as clonal sharing between patients, additional experiments are needed. Second, we focused on repertoire abnormalities in heavy chains based on their central roles in antigen recognition. The characterization of light chains, as well as the combination of heavy and light chains with single-cell analysis, would further clarify the repertoire characteristics in diseases.

In summary, our large-scale BCR repertoire analysis provided an overview of B cell abnormalities in IMDs.

This analysis exemplifies the usefulness of multi-omics human data for disentangling the complexity of cellular biology.

## Data Availability

BCR data used in this study will be available at the National Bioscience Database Center (NBDC) Human Database (https://humandbs.biosciencedbc.jp/en/) at the time of publication.

## List of Supplementary Materials

Fig. S1 to S5 and Table S5 in Supplementary Information (PDF file). Table S1 to S4 and Table S6 in Supplementary Tables (Excel file).

## Acknowledgements

The super-computing resource was provided by Human Genome Center, Institute of Medical Sciences, The University of Tokyo (http://sc.hgc.jp/shirokane.html).

## Author Contributions

MO conducted bioinformatics analysis with the help of YN and KI. MN contributed to the collection and management of clinical information. MO, MN, YN, SK, HH, RY, TI, YA, HS, and NB managed and contributed to sample collection, cell sorting, RNA sequencing and whole genome sequencing. HH, YA, and TI contributed to QC of the RNA-seq data. NB performed BCR-seq experiment. TO, KY and KF designed and managed the project. YT contributed to critical reading and revision of the manuscript. MO and KF designed the study and wrote the manuscript with contributions from all authors on the final version of the manuscript.

## Data and Code Availability

BCR data used in this study is deposited at the National Bioscience Database Center (NBDC) Human Database (https://humandbs.biosciencedbc.jp/en/) with study ID of JGAS000485 and will be available upon acceptance. We used publicly available software for the analyses. Custom code is available from the corresponding authors upon reasonable request.

## References

1. Kundaje A, Meuleman W, Ernst J, et al. Integrative analysis of 111 reference human epigenomes. Nature 2015;518(7539):317–30.

2. Ota M, Nagafuchi Y, Hatano H, et al. Dynamic landscape of immune cell-specific gene regulation in immune-mediated diseases. Cell 2021;184(11):3006-21.e17.

3. Mitka M. Treatment for lupus, first in 50 years, offers modest benefits, hope to patients. Jama 2011;305(17):1754–5.

4. Edwards JC, Szczepanski L, Szechinski J, et al. Efficacy of B-cell-targeted therapy with rituximab in patients with rheumatoid arthritis. The New England journal of medicine 2004;350(25):2572–81.

5. van Kempen TS, Wenink MH, Leijten EF, et al. Perception of self: distinguishing autoimmunity from autoinflammation. Nat Rev Rheumatol 2015;11(8):483–92.

6. Elsner RA, Shlomchik MJ. Germinal Center and Extrafollicular B Cell Responses in Vaccination, Immunity, and Autoimmunity. Immunity 2020;53(6):1136–50.

7. Jenks SA, Cashman KS, Zumaquero E, et al. Distinct Effector B Cells Induced by Unregulated Toll-like Receptor 7 Contribute to Pathogenic Responses in Systemic Lupus Erythematosus. Immunity 2018;49(4):725-39.e6.

8. Tipton CM, Fucile CF, Darce J, et al. Diversity, cellular origin and autoreactivity of antibody-secreting cell population expansions in acute systemic lupus erythematosus. Nat Immunol 2015;16(7):755–65.

9. Brown GJ, Cañete PF, Wang H, et al. TLR7 gain-of-function genetic variation causes human lupus. Nature 2022;605(7909):349–56.

10. Glass DR, Tsai AG, Oliveria JP, et al. An Integrated Multi-omic Single-Cell Atlas of Human B Cell Identity. Immunity 2020;53(1):217-32.e5.

11. Miqueu P, Guillet M, Degauque N, et al. Statistical analysis of CDR3 length distributions for the assessment of T and B cell repertoire biases. Molecular immunology 2007;44(6):1057–64.

12. Gomez-Tourino I, Kamra Y, Baptista R, et al. T cell receptor β-chains display abnormal shortening and repertoire sharing in type 1 diabetes. Nat Commun 2017;8(1):1792.

13. Palanichamy A, Bauer JW, Yalavarthi S, et al. Neutrophil-mediated IFN activation in the bone marrow alters B cell development in human and murine systemic lupus erythematosus. Journal of immunology (Baltimore, Md : 1950) 2014;192(3):906–18.

14. Mroczek ES, Ippolito GC, Rogosch T, et al. Differences in the composition of the human antibody repertoire by B cell subsets in the blood. Front Immunol 2014;5:96.

15. Kaplinsky J, Li A, Sun A, et al. Antibody repertoire deep sequencing reveals antigen-independent selection in maturing B cells. Proceedings of the National Academy of Sciences of the United States of America 2014;111(25):E2622–9.

16. Bashford-Rogers RJM, Bergamaschi L, McKinney EF, et al. Analysis of the B cell receptor repertoire in six immune-mediated diseases. Nature 2019;574(7776):122–26.

17. Demaison C, David D, Fautrel B, et al. V(H) gene-family representation in peripheral activated B cells from systemic lupus erythematosus (SLE) patients. Clinical and experimental immunology 1996;104(3):439–45.

18. King HW, Orban N, Riches JC, et al. Single-cell analysis of human B cell maturation predicts how antibody class switching shapes selection dynamics. Sci Immunol 2021;6(56).

19. Sankar K, Hoi KH, Hötzel I. Dynamics of heavy chain junctional length biases in antibody repertoires. Commun Biol 2020;3(1):207.

20. William J, Euler C, Christensen S, et al. Evolution of autoantibody responses via somatic hypermutation outside of germinal centers. Science (New York, NY) 2002;297(5589):2066–70.

21. Ghraichy M, Galson JD, Kovaltsuk A, et al. Maturation of the Human Immunoglobulin Heavy Chain Repertoire With Age. Front Immunol 2020;11:1734.

22. Rao DA, Gurish MF, Marshall JL, et al. Pathologically expanded peripheral T helper cell subset drives B cells in rheumatoid arthritis. Nature 2017;542(7639):110–14.

23. Takeshima Y, Iwasaki Y, Nakano M, et al. Immune cell multiomics analysis reveals contribution of oxidative phosphorylation to B-cell functions and organ damage of lupus. Ann Rheum Dis 2022; 81(6):845–853.

24. Bocharnikov AV, Keegan J, Wacleche VS, et al. PD-1hiCXCR5-T peripheral helper cells promote B cell responses in lupus via MAF and IL-21. JCI Insight 2019;4(20).

25. Makiyama A, Chiba A, Noto D, et al. Expanded circulating peripheral helper T cells in systemic lupus erythematosus: association with disease activity and B cell differentiation. Rheumatology (Oxford, England) 2019;58(10):1861–69.

26. Kenderes KJ, Levack RC, Papillion AM, et al. T-Bet(+) IgM Memory Cells Generate Multi-lineage Effector B Cells. Cell Rep 2018;24(4):824-37.e3.

27. Rubtsova K, Rubtsov AV, Thurman JM, et al. B cells expressing the transcription factor T-bet drive lupus-like autoimmunity. J Clin Invest 2017;127(4):1392–404.

28. Reed JH, Jackson J, Christ D, et al. Clonal redemption of autoantibodies by somatic hypermutation away from self-reactivity during human immunization. The Journal of experimental medicine 2016;213(7):1255–65.

29. Schickel J-N, Glauzy S, Ng Y-S, et al. Self-reactive VH4-34-expressing IgG B cells recognize commensal bacteria. The Journal of experimental medicine 2017;214(7):1991–2003.

30. Banchereau J, Pascual V. Type I interferon in systemic lupus erythematosus and other autoimmune diseases. Immunity 2006;25(3):383–92.

31. Tobón GJ, Izquierdo JH, Cañas CA. B lymphocytes: development, tolerance, and their role in autoimmunity-focus on systemic lupus erythematosus. Autoimmune Dis 2013;2013:827254.

32. Liu S, Hou XL, Sui WG, et al. Direct measurement of B-cell receptor repertoire’s composition and variation in systemic lupus erythematosus. Genes Immun 2017;18(1):22–27.

33. Meffre E, Milili M, Blanco-Betancourt C, et al. Immunoglobulin heavy chain expression shapes the B cell receptor repertoire in human B cell development. J Clin Invest 2001;108(6):879–86.

34. DeWitt WS, Lindau P, Snyder TM, et al. A Public Database of Memory and Naive B-Cell Receptor Sequences. PLoS One 2016;11(8):e0160853.

35. Hou X, Zeng P, Zhang X, et al. Shorter TCR β-Chains Are Highly Enriched During Thymic Selection and Antigen-Driven Selection. Front Immunol 2019;10:299.

36. Yurasov S, Wardemann H, Hammersen J, et al. Defective B cell tolerance checkpoints in systemic lupus erythematosus. The Journal of experimental medicine 2005;201(5):703–11.

37. Cappione A, 3rd, Anolik JH, Pugh-Bernard A, et al. Germinal center exclusion of autoreactive B cells is defective in human systemic lupus erythematosus. J Clin Invest 2005;115(11):3205–16.

38. Huang W, Quach TD, Dascalu C, et al. Belimumab promotes negative selection of activated autoreactive B cells in systemic lupus erythematosus patients. JCI Insight 2018;3(17).

