## Supplementary information for "Multimodal repertoire analysis unveils B cell biology in immune-mediated diseases"

**Online Supplementary Information**

### MATERIALS AND METHODS

#### Criteria for patient inclusion/exclusion

The SLE cohort was comprised of patients who had attended the Department of Allergy and Rheumatology at the University of Tokyo Hospital, Division of Rheumatic Diseases at National Center for Global Health and Medicine or Immuno-Rheumatology Center at St. Luke's International Hospital, who met the 1997 revised version of ACR SLE criteria [1]. Of the enrolled 136 patients, 88 cases had active disease with SLEDAI-2K ≥ 4. The other cases were inactive at the time of blood withdrawal. Patients administered cyclophosphamide or rituximab within one year or taking ≥ 21 mg/day prednisolone were excluded from this analysis. For 22 patients, belimumab was initiated after the collection of the samples, and samples after a half-year treatment were also collected. To assess the drug's effect, cases with concomitant prednisolone increase ≥ 5 mg/day or concomitant addition of other immunosuppressants were excluded.

The SSc cohort was comprised of patients who had attended the Department of Allergy and Rheumatology at the University of Tokyo Hospital or Department of Rheumatology at Tokyo Metropolitan Komagome Hospital, who met the 2013 American College of Rheumatology (ACR)/ European League Against Rheumatism (EULAR) classification criteria for Systemic Sclerosis [2]. Patients taking ≥ 21 mg/day of prednisolone were excluded from this analysis.

The IIM cohort was comprised of patients who had attended to the Department of Allergy and Rheumatology at the University of Tokyo Hospital or Division of Rheumatology at the Jikei University Hospital. They met standards defined by the Bohan and Peter criteria [3 4], or the European Neuromuscular Center criteria [5], or the Sontheimer criteria [6] or the Griggs criteria [7]. Patients taking ≥ 12 mg/day prednisolone were excluded. Of the 85 patients, 36 cases had active disease and required subsequent initiation or an increase of immunomodulatory drugs.

The inclusion criteria for HC were people with no apparent co-morbidities, no direct family history of autoimmune diseases and no use of prescription drugs or supplements. Age and sex were matched with the patient cohort as much as possible. For 30 HC cases, only naïve B cells among 5 B cell subsets were collected and utilized for the analysis. For the remaining 107 cases, 5 B cell subsets were collected.

Patient inclusion/exclusion criteria for the other diseases were described in detail in our first report [8].

Samples were collected from the University of Tokyo, the Jikei University School of Medicine, St. Luke's International Hospital, National Center for Global Health, and Medicine and Tokyo Metropolitan

Komagome Hospital with approval by the Ethics Committees at each site. Written informed consent was obtained from all subjects enrolled.

**RNA-seq experiments**

We followed the same protocol that we reported previously [8]. Briefly, peripheral blood mononuclear cells (PBMCs) were isolated by density gradient separation with Ficoll-Paque (GE Healthcare) immediately after blood drawing. After lysis of erythrocytes, up to 26 immune cell subsets were collected with purity > 99% using a MoFlo XDP instrument (Beckman Coulter) or a 14-colour cell sorter BD FACSAria Fusion (BD Biosciences). We intended to collect a minimum of 1,000 cells (up to 5,000 cells) per subset. Libraries for RNA-seq were prepared using SMART-seq v4 Ultra Low Input RNA Kits (Takara Bio). Samples failing any of the quality control steps (RNA quality and quantity, PCR amplification, library fragment size) were eliminated from further downstream steps. Libraries were sequenced on the HiSeq 2500 Illumina platform to obtain 100-bp paired-end reads with HiSeq SBS Kit v4 (Illumina).

**Mapping of the transcriptome**

We followed the same protocol we reported previously [8]. Briefly, adaptor sequences were trimmed using cutadapt (v1.16) and 3′- ends with low-quality bases (Phred quality score < 20) were trimmed using the fastx-toolkit (v0.0.14). Reads containing more than 20% low-quality bases were removed. Subsequently, reads were aligned against the GRCh38 reference sequence using STAR (v2.5.3) [9] in two-pass mode with GENCODE, version 27 annotations [10]. A median of 10.1 million paired fragments were uniquely mapped per sample and utilized for the subsequent analysis. Expression was quantified using HTSeq (v 0.11.2) [11].

**Mapping and QC of BCR sequences from RNA-seq data**

We utilized MiXCR software (version 3.0.13) [12] for BCR mapping. With FASTQ files after adapter and quality trimming, we utilized the “analyze shotgun” option with parameters “-s hsa", “--starting-material rna” and “--receptor-type bcr”. After alignment and assembly of clonotypes based on the CDR3 sequence, the full BCR sequence was assembled with the “--contig-assembly” parameter. We set “specificMutationProbability” parameter to $2 \times 10^{-3}$. Only productive heavy-chain sequences were kept for the analysis unless specified. For all analyses, samples with more than 500 unique CDR-H3 sequences were used for the analysis. For each sample, we treated each unique CDR3 sequence as a single event, regardless of read depth. For mapping non-productive sequences, we raised the "specificMutationProbability" parameter to $8 \times 10^{-3}$ in order to reduce the amount of productive sequences with error or mutations counted as non-productive sequences.

**BCR-seq experiments**

We utilized residual RNA samples after RNA-seq for BCR-targeted sequence. We chose 20 samples which had the wide range in the number of BCR-mapped reads and sufficient volume of residual RNA: 5 samples from naïve B cells, 6 samples from USM B and 9 samples from plasmablasts.

BCR region amplification and library preparation was performed with SMARTer Human BCR IgG IgM H/K/L Profiling Kit (Takara Bio). Heavy chain IgM sequences were amplified for naïve B cells and USM B, and heavy chain IgG sequences were amplified for plasmablasts. Libraries were sequenced on the MiSeq Illumina platform to obtain 300-bp paired-end reads. We utilized the same version of MiXCR software (version 3.0.13) as RNA-seq data analysis for BCR-seq data analysis. All samples were used for the comparison with RNA-seq.

**Principal component analysis based on VDJ gene usage**

All B cell samples used in this study except those samples after belimumab treatment were utilized for PCA analysis. For each sample, each VDJ gene usage was calculated by dividing the number of clonotypes using that gene by the total number of clonotypes in that sample. We retained genes that were used more than 0.5% by 10% or more of the samples. Each gene usage was scaled to mean = 0 and variance = 1, and PCA was performed with prcomp function in R.

To project samples after belimumab treatment to the same PCA space described above, for each VDJ gene, we centered the mean and standardized the variance with regards to those calculated among all the other B cell samples above. Then, this normalized VDJ gene usage matrix and the PC loadings of each gene calculated in the above analysis were utilized for calculating PC scores for samples after belimumab treatment.

We assessed the disease effects on PC scores by linear regression analysis comparing each disease sample versus HC samples in each subset. Treatment effects on PC scores were assessed by the linear mixed model comparing samples before and after treatment, with individuals treated as random effects. The linear mixed model was fit with lme4 package.

**Calculation of the Repertoire Naïveness Score**

For scoring, we only included VDJ genes that were used by more than an average of 0.1% of the clonotypes in each of the 5 subsets. At first, each VDJ gene usage was scaled to mean = 0 and variance = 1 among the 5 B cell subsets of HCs. Then, the effect sizes of VDJ genes were estimated by comparing their usage between naïve B and USM B among HC samples with a linear mixed model, with individuals treated as random effects. Only genes that attained Bonferroni-adjusted p values < 0.05 in this comparison were utilized for the subsequent scoring. In addition, genes whose usage showed substantial correlation among 5 B cell subsets of HCs ($|\beta| > 0.4$ in the linear mixed model) were pruned to keep the most significantly different ones in the comparison of USM B and naïve B. As a result, 55 genes (35 V genes, 16 D genes and 4 J genes) were retained for the scoring (Table S3).

For each sample, VDJ gene usage was mean-centered and variance-standardized with regards to those calculated among the 5 B cell subsets of HCs. Then, the following formula was applied for calculating the RN score:

$$RNS_i = \sum_{k=1}^{35} \hat{\beta}_{V_k} \times U_{V_{ki}} + \sum_{k=1}^{16} \hat{\beta}_{D_k} \times U_{D_{ki}} + \sum_{k=1}^{4} \hat{\beta}_{J_k} \times U_{J_{ki}}$$

where $\hat{\beta}_{V_k}$ corresponded to the effect size of the $V_k$ gene usage as calculated above, and $U_{V_{ki}}$ corresponded to the normalized usage of the $V_k$ gene in individual $i$.

**Calculation of gene set enrichment score**

We utilized the GSVA package [13] to calculate the gene set enrichment score for samples. After TMM normalization [14] of count data, conversion to log count per million values and batch effect removal with the ComBat function from the sva package [15], the "gsva" function was utilized for calculating the gene set enrichment score for the gene sets in Table S6. The Tph1 score was calculated using the 54 genes that were significantly upregulated in PD-1$^{hi}$ CD4$^{+}$ T cells compared with PD-1$^{-}$ populations in the original article [16] and normalized expression values of Th1 cells. The IFN score was calculated using 97 genes in HALLMARK_INTERFERON_ALPHA_RESPONSE gene set in MSigDB [17].

**Association analysis with gene expression**

The association of naïve B cell CDR-H3 length with gene expression in naïve B cells was assessed in each disease at first for reducing the confounding of inter-disease differences in gene expression-CDR-H3 length association. For this step, gene expression data after filtering of low expressed genes was assessed for the association with CDR-H3 length using the limma [18] package with voom transformation. Known batch effects and age were treated as covariates. Then, resultant effect sizes of each comparison and their standard deviations were utilized for fixed effect meta-analysis with Metasoft (version 2.0.1) software [19].

For the assessment of association of RN score and gene expression in each immune cell subset, we also utilized limma with voom transformation. The RN score was scaled to mean = 0 and variance = 1 and known batch effects and age were treated as covariates. In this comparison, genes fulfilling FDR < 0.05 and |logFC| > 0.1 were treated as significantly associated genes.

**Gene set enrichment analysis**

We tested the enrichment of CDR-H3 length-associated genes that passed a Bonferroni significance threshold in fixed-effect meta-analysis to hallmark gene sets by MsigDB [17] using the clusterProfiler package [20].

**Differential usage analysis**

For the assessment of disease effects on VDJ gene usage in each subset, we kept genes that were used more than 0.5% by 10% or more of the samples. For each disease, gene usage of the disease and HC samples were scaled to mean = 0 and variance = 1 and tested for the disease effect on gene usage by linear regression with age and known batch effects as covariates.

**Mediation analysis**

The relationships among IFN scores, Tph1 scores and RN scores in plasmablasts were tested among SLE patients by utilizing the classical mediation analysis framework [21]. The IFN score was treated as the independent variable, the Tph1 score was treated as the mediator and the RN score was treated as the outcome variable. P values were calculated via 10000-time bootstrapping using mediation package in R.

**Calculation of V gene mutation frequency**

For the assessment of gene mutation frequency, we quantified per-base mutation frequency in the heavy-chain V gene except for the CDR3 region, that is, in FR1, CDR1, FR2, CDR2 and FR3 regions. As MiXCR assembles the sequence of these regions after alignment and assembly of clonotypes based on CDR3 sequence, only a portion of the nucleotides is actually sequenced in these regions. We counted the number of mutations in these regions in each individual and divided it by the total number of sequenced nucleotides in these regions in the individual to infer the V gene mutation frequency.

**Comparison with the data from the other cohort**

The BCR repertoire sequencing data of sorted B cells from human tonsil [22] was downloaded from ArrayExpress with the accession number E-MTAB-8999.

Gene expression data of PD-1$^{hi}$ and PD-1$^{−}$ T cells [16] were downloaded from the ImmPort repository with the accession number SDY939. We performed the differential expression analysis between (memoryCD4$^{+}$PD-1$^{hi}$CXCR5$^{-}$MHCII$^{-}$ICOS$^{+}$ + memoryCD4$^{+}$PD-1$^{hi}$CXCR5$^{-}$MHCII$^{+}$ICOS$^{+}$) versus memoryCD4$^{+}$PD-1$^{-}$CXCR5$^{-}$ populations to obtain "PD-1$^{hi}$ versus PD-1$^{−}$ logFC".

To validate the reproducibility of the correlation between Tph1 transcriptome signature and RN score in plasmablasts, we utilized peripheral blood Th1 cells and plasmablast RNA-seq data of Japanese SLE patients and HCs that we had obtained previously [23]. We excluded the overlapped participants with this study and this dataset consisted of 30 SLE patients and 26 HCs.

**Comparison of the frequency of IGHV4-34 clonotypes with nonmutated AVY/NHS motifs**

We counted the number of IGHV4-34 clonotypes with or without mutations in the AVY motif in the FWR1 region or the NHS motif in the CDR2 region. Only clonotypes sequenced at these positions were included in the analysis. As the number of IGHV4-34 clonotypes was not large in one sample, we aggregated the IGHV4-34$^{+}$ clonotypes in each comparison group (e.g., SLE plasmablasts, HC plasmablasts, etc.), and compared the cumulative number of mutated and nonmutated clonotypes between groups by Fisher's exact test.

**Data visualization and statistical analysis**

The statistical test performed are indicated in the figure legends or Methods. Throughout the analyses, multiple test correction was performed with Benjamini-Hochberg procedure to obtain FDRs unless otherwise indicated. In boxplots, the center lines indicate the median, the box limits indicate the interquartile range (IQR), the whiskers reflect the maximum and minimum values no further than 1.5 × IQR from the hinge and the points indicate outliers.

### Supplementary Figures

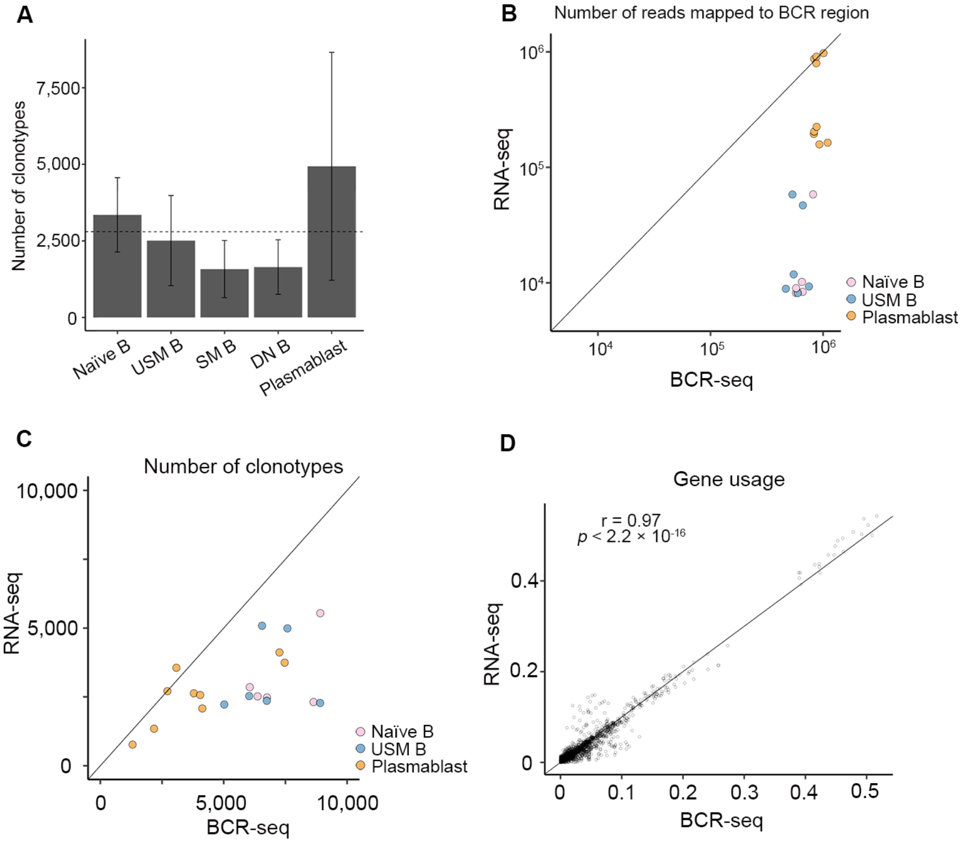


**Fig. S1. BCR repertoire characteristics and comparison between RNA-seq and BCR-seq, related to Fig. 1**

**(A)** Mean number of clonotypes identified in each subset in our cohort. Bars indicate 2SD. Dotted line indicates the mean number of identified clonotypes among 5 subsets. **(B-D)** Comparison of total number of clonotype counts which reflect the effective read counts (B), number of identified clonotypes (C), and VDJ gene usage (D) between the results from RNA-seq experiment and BCR-seq experiment. Solid lines indicate y = x.

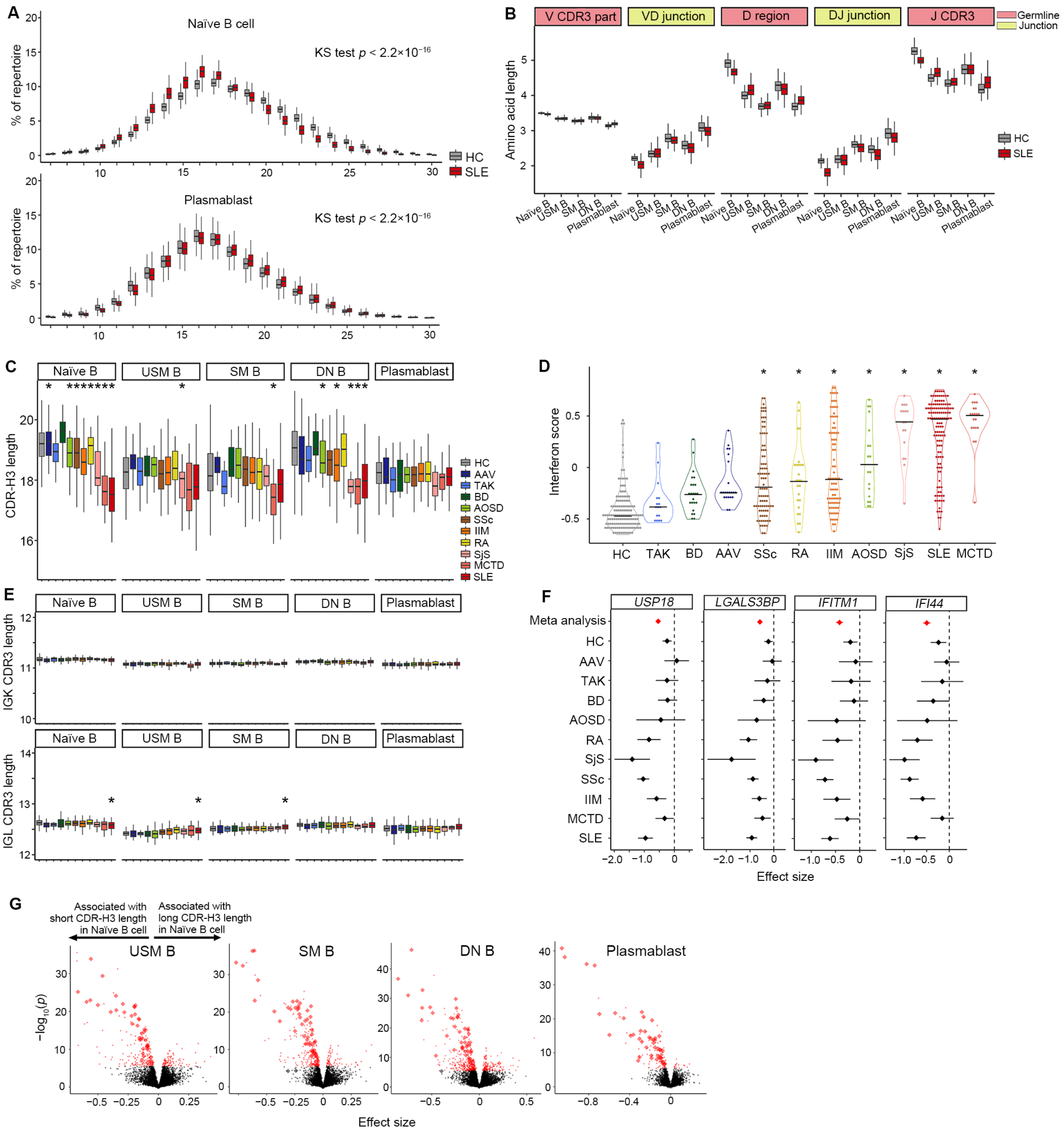

**Fig. S2. Association of CDR-H3 length with IMDs and gene expression, related to Fig. 2**

**(A)** Comparison of distribution of CDR-H3 lengths in SLE and HC in naïve B cells and plasmablasts. KS test, Kolmogorov-Smirnov test. **(B)** Comparison of lengths of sections that constitute the CDR-H3 region, related to Fig. 2B. **(C)** Comparison of CDR-H3 lengths between IMDs and HC among non-productive clonotypes. Differences between HC and each disease were tested with linear regression adjusting for age and batch effects. *, $p < 0.05/50$. **(D)** Comparison of IFN scores in naïve B cells among IMD and HC. Median values are marked with black line. Differences between HC and each disease were tested with linear regression adjusting for age and batch effects. *, $p < 0.05/10$. **(E)** Comparison of light chain CDR3 length between IMD and HC. Differences between HC and each disease were tested with linear regression adjusting for age and batch effects. *, $p < 0.05/50$. **(F)** Effect sizes for the associations of labeled genes and CDR-H3 lengths in each disease as well as fixed effect meta-analysis. Bars indicate 95% CI. **(G)** Association of gene expression in 4 B cell subsets with naïve B CDR-H3 length, related to Fig. 2C. Fixed effect meta-analysis *p* values and effect sizes are plotted. Genes attained Bonferroni-adjusted $p < 0.05$ are marked red. Genes in "HALLMARK_INTERFERON_ALPHA_RESPONSE" gene set are marked as diamonds.

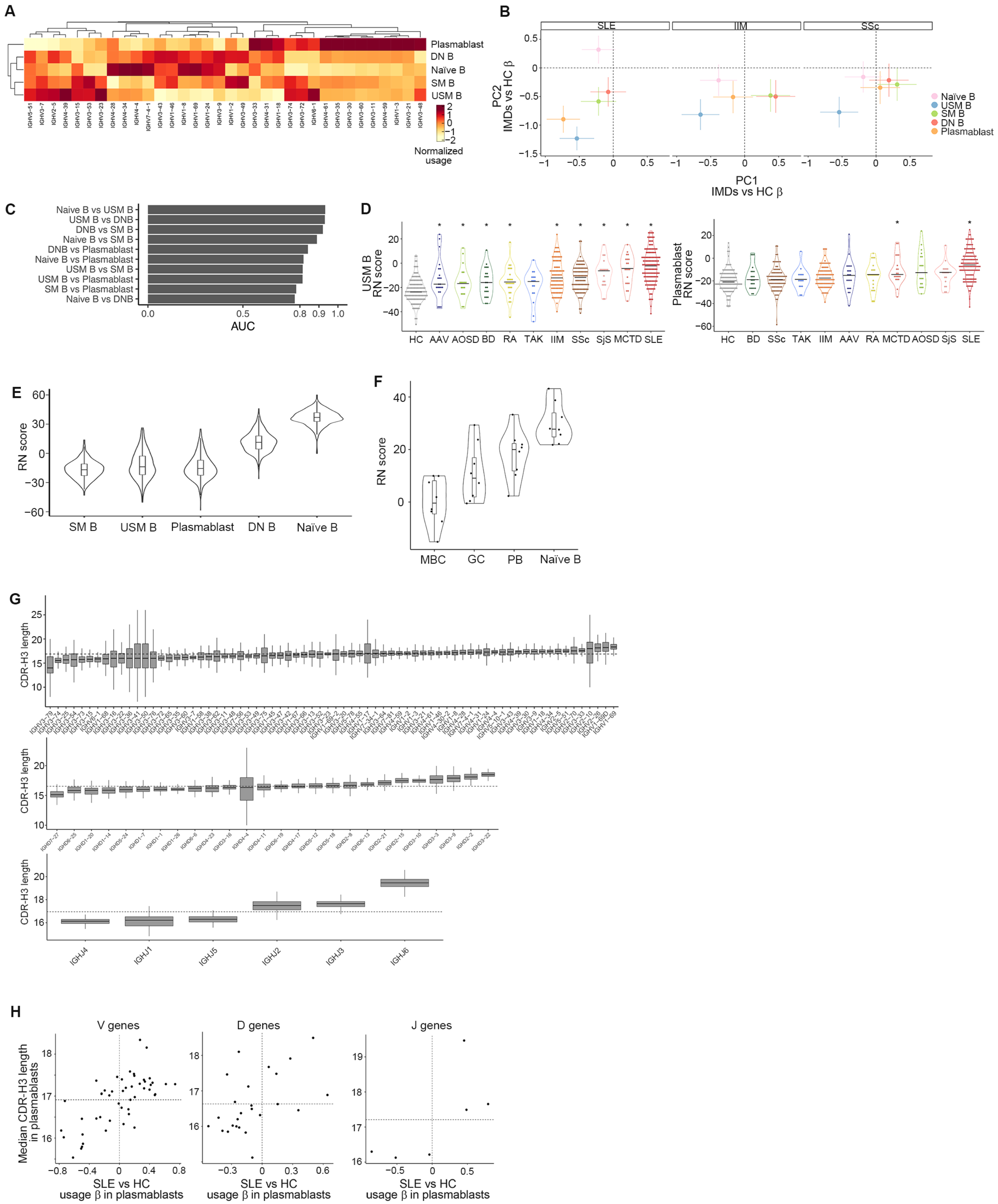

A
Plasmablast
DN B
Naïve B
SM B
USM B
Normalized usage
B
SLE
IIM
SSc
PC2 IMDs vs HC β
PC1 IMDs vs HC β
Naïve B
USM B
SM B
DN B
Plasmablast
C
Naive B vs USM B
USM B vs DNB
DNB vs SM B
Naive B vs SM B
DNB vs Plasmablast
Naive B vs Plasmablast
USM B vs SM B
USM B vs Plasmablast
SM B vs Plasmablast
Naive B vs DNB
AUC
D
USM B RN score
HC AAV AOSD BD RA TAK IIM SSc SjS MCTD SLE
Plasmablast RN score
HC BD SSc TAK IIM AAV RA MCTD AOSD SjS SLE
E
RN score
SM B USM B Plasmablast DN B Naïve B
F
RN score
MBC GC PB Naïve B
G
CDR-H3 length
IGHJ4 IGHJ1 IGHJ5 IGHJ2 IGHJ3 IGHJ6
H
V genes
D genes
J genes
Median CDR-H3 length in plasmablasts
SLE vs HC usage β in plasmablasts

**Fig. S3. Comparison of gene usage patterns between IMDs and evaluation of the Repertoire Naïveness score, related to Fig. 3**

**(A)** Comparison of mean V gene usage among 5 B cell subsets in HC. Genes that are utilized at least 1% in any of 5 subsets are shown. For comparison, gene usages were Z score normalized column-wise and hierarchically clustered. **(B)** Disease effects on PC scores generated by linear regression analysis: comparison of each disease sample versus HC samples. Bars indicate 95% CI. **(C)** Comparison of gene usage-based scoring methods. Weights of VDJ genes used for scoring were calculated in 10 different ways (y axis) based on the gene usage difference between B cell subsets in HC. The performance of each scoring method was evaluated by AUC for distinguishing SLE patients from HC using the scores in 5 B cell subsets by fitting the logistic regression model. **(D)** Comparison of RN scores determined for 5 B cell subsets among all participants in our cohort. **(E)** Comparison of RN scores among IMDs in USM B (left) and plasmablasts (right). Black bars indicate median values. Differences between HC and each disease were tested with linear regression adjusting for age and batch effects. *, $p < 0.05/10$. **(F)** Comparison of RN scores among previously reported BCR-seq data of sorted B cells from human tonsil. MBC, memory B cell. GC, germinal center B cell. PB, plasmablast. Naïve, naïve B cell. **(G)** Distribution of median CDR-H3 lengths of clonotypes utilizing each V (top), D (middle), and J (bottom) gene in plasmablasts among HC. Dotted lines indicate the median CDR-H3 length among all clonotypes. **(H)** Comparison of the effect size estimates of SLE versus HC gene usage comparison in plasmablasts (x-axis) and median CDR-H3 length of each gene among HC in plasmablasts (y-axis).

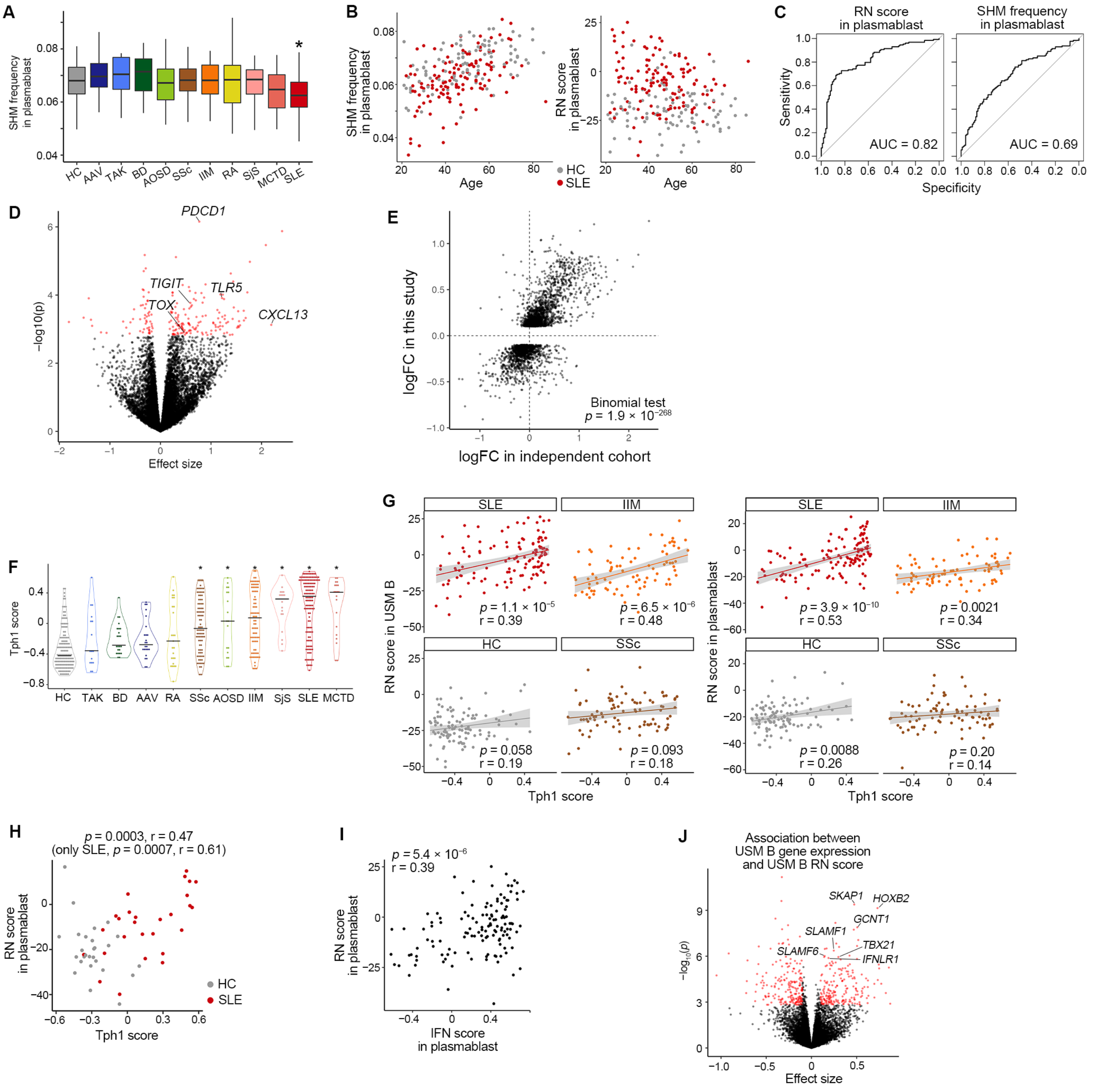

**Fig. S4. RN score-associated gene expression profiles and their reproducibility in the independent cohort, related to Fig. 4**

**(A)** Comparison of SHM frequencies in the V gene region of plasmablasts among IMDs. Differences between HC and each disease were tested with linear regression adjusting for age and batch effects. *, $p < 0.05/10$. **(B)** Association of SHM frequencies (left) and RN scores of plasmablasts (right) with age. **(C)**

ROC curve for classifying SLE patients from HC using RN score in plasmablasts (left) or SHM frequencies (right) in plasmablasts. **(D)** Association of gene expression in Th1 cells and RN score among SLE patients in the independent cohort. RN score was calculated in plasmablasts. Significantly associated genes are colored red. **(E)** Log fold change in Th1 expression versus RN score comparison in two different cohorts. Each dot corresponds to each gene. They showed high concordance between SLE patients in this study and the independent SLE cohort. RN score was calculated in plasmablasts. **(F)** Comparison of Tph1 scores. Differences between HC and each disease were tested with linear regression adjusting for age and batch effects. *, $p < 0.05/10$. **(G)** Correlation between Tph1 scores and RN scores in USM B (left) or plasmablasts (right). Pearson's r and its significance are provided. **(H)** Correlation between Tph1 scores and RN scores in plasmablasts in the independent cohort. **(I)** Correlation between IFN scores and RN scores in plasmablasts among SLE patients. **(J)** Association of gene expression in USM B cells and RN scores in USM B. Significantly associated genes are marked red.

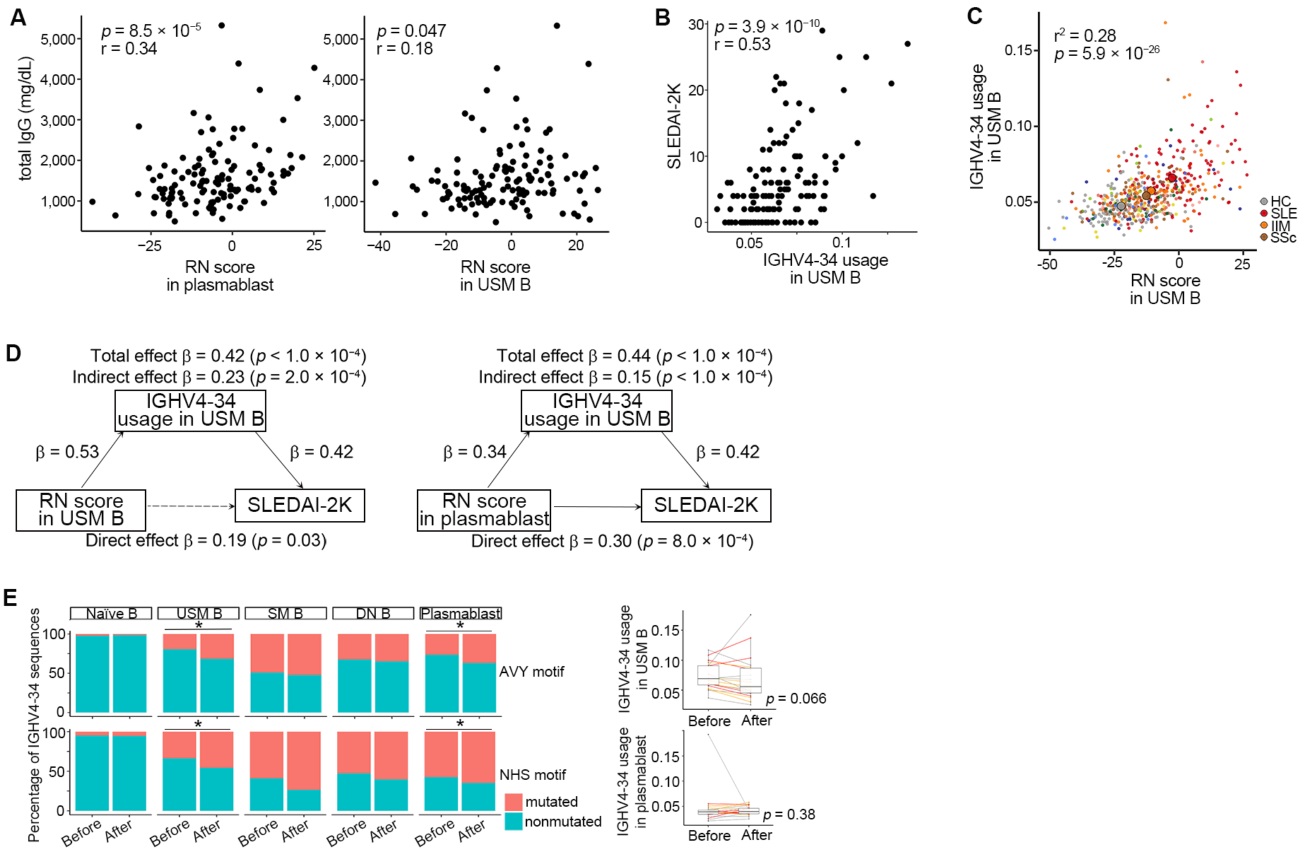


**Fig. S5. Comparison of RN score, VH4-34 motifs and isotype frequencies, related to Fig. 5**

**(A)** Correlation between RN scores and serum total IgG concentrations. Pearson's r and its significance are provided. **(B)** Correlation between IGHV4-34 usage in USM B and SLEDAI-2K. Pearson's r and its significance are provided. **(C)** Correlation between RN scores and IGHV4-34 usage in USM B. $R^2$ and $p$ values were calculated with linear regression model. Mean values among HC, SLE, SSc, and IIM are marked with large points. The color of each point indicates the clinical diagnosis as illustrated in Fig. 1A. **(D)** Mediation model representing the relationships among RN scores in USM B (left) or plasmablasts (right), IGHV4-34 usage in USM B and SLEDAI-2K among SLE patients. **(E)** Comparison of the frequencies of clonotypes with mutations in AVY/NHS motifs among IGHV4-34 clonotypes (left) and IGHV4-34 usage (right) before and after treatment with belimumab. For motif comparison, Fisher's exact test comparing the number of mutated/nonmutated clonotypes before and after treatment samples was performed in each subset; *, $p < 0.01$.

**Table S5. Abbreviations for immune cell subsets, Related to Figure 4**

| Abbreviation | Subset name |
|---|---|
| CD4 | CD4 T cells |
| Naïve CD4 | Naïve CD4 T cells |
| Mem CD4 | Memory CD4 T cells |
| Th1 | T helper 1 cells |
| Th2 | T helper 2 cells |
| Th17 | T helper 17 cells |
| Tfh | T follicular helper cells |
| Fr. II eTreg | Fraction II effector regulatory T cells |
| Fr. I nTreg | Fraction I naïve regulatory T cells |
| Fr. III T | Fraction III non-regulatory T cells |
| CD8 | CD8 T cells |
| Naïve CD8 | Naïve CD8 T cells |
| TEMRA CD8 | CD8+ T effector memory CD45RA+ cells |
| EM CD8 | Effector Memory CD8 T cells |
| CM CD8 | Central Memory CD8 T cells |
| B | B cells |
| Naïve B | Naïve B cells |
| USM B | Unswitched memory B cells |
| SM B | Switched memory B cells |
| DN B | Double Negative B cells |
| Plasmablast | Plasmablasts |
| NK | Natural Killer cells |
| Mono | Monocytes |
| CD16p Mono | CD16 positive monocytes |
| NC Mono | Non-classical monocytes |
| Int Mono | Intermediate monocytes |
| CL Mono | Classical monocytes |
| DC | Dendric cells |
| mDC | Myeloid dendritic cells |
| pDC | Plasmacytoid dendritic cells |
| Neu | Neutrophils |
| LDG | Low-Density Granulocytes |

**Supplementary References**


1. Hochberg MC. Updating the American College of Rheumatology revised criteria for the classification of systemic lupus erythematosus. *Arthritis Rheum* 1997;**40**(9):1725.
2. van den Hoogen F, Khanna D, Fransen J, et al. 2013 classification criteria for systemic sclerosis: an American College of Rheumatology/European League against Rheumatism collaborative initiative. *Arthritis Rheum* 2013;**65**(11):2737-47.
3. Bohan A, Peter JB. Polymyositis and dermatomyositis (first of two parts). *The New England journal of medicine* 1975;**292**(7):344-7.
4. Bohan A, Peter JB. Polymyositis and dermatomyositis (second of two parts). *The New England journal of medicine* 1975;**292**(8):403-7.
5. Hoogendijk JE, Amato AA, Lecky BR, et al. 119th ENMC international workshop: trial design in adult idiopathic inflammatory myopathies, with the exception of inclusion body myositis, 10-12 October 2003, Naarden, The Netherlands. Neuromuscul Disord. England, 2004:337-45.
6. Sontheimer RD. Would a new name hasten the acceptance of amyopathic dermatomyositis (dermatomyositis sine myositis) as a distinctive subset within the idiopathic inflammatory dermatomyopathies spectrum of clinical illness? *J Am Acad Dermatol* 2002;**46**(4):626-36.
7. Griggs RC, Askanas V, DiMauro S, et al. Inclusion body myositis and myopathies. *Ann Neurol* 1995;**38**(5):705-13.
8. Ota M, Nagafuchi Y, Hatano H, et al. Dynamic landscape of immune cell-specific gene regulation in immune-mediated diseases. *Cell* 2021;**184**(11):3006-21.e17.
9. Dobin A, Davis CA, Schlesinger F, et al. STAR: ultrafast universal RNA-seq aligner. *Bioinformatics (Oxford, England)* 2013;**29**(1):15-21.
10. Frankish A, Diekhans M, Ferreira AM, et al. GENCODE reference annotation for the human and mouse genomes. *Nucleic acids research* 2019;**47**(D1):D766-d73.
11. Anders S, Pyl PT, Huber W. HTSeq--a Python framework to work with high-throughput sequencing data. *Bioinformatics (Oxford, England)* 2015;**31**(2):166-9.
12. Bolotin DA, Poslavsky S, Mitrophanov I, et al. MiXCR: software for comprehensive adaptive immunity profiling. *Nat Methods* 2015;**12**(5):380-1.
13. Hänzelmann S, Castelo R, Guinney J. GSVA: gene set variation analysis for microarray and RNA-seq data. *BMC Bioinformatics* 2013;**14**:7.
14. Robinson MD, McCarthy DJ, Smyth GK. edgeR: a Bioconductor package for differential expression analysis of digital gene expression data. *Bioinformatics (Oxford, England)* 2010;**26**(1):139-40.
15. Leek JT, Johnson WE, Parker HS, et al. The sva package for removing batch effects and other unwanted variation in high-throughput experiments. *Bioinformatics (Oxford, England)* 2012;**28**(6):882-3.
16. Rao DA, Gurish MF, Marshall JL, et al. Pathologically expanded peripheral T helper cell subset drives B cells in rheumatoid arthritis. *Nature* 2017;**542**(7639):110-14.

17. Liberzon A, Birger C, Thorvaldsdóttir H, et al. The Molecular Signatures Database (MSigDB) hallmark gene set collection. *Cell Syst* 2015;**1**(6):417-25.

18. Ritchie ME, Phipson B, Wu D, et al. limma powers differential expression analyses for RNA-sequencing and microarray studies. *Nucleic acids research* 2015;**43**(7):e47.

19. Han B, Eskin E. Interpreting meta-analyses of genome-wide association studies. *PLoS genetics* 2012;**8**(3):e1002555.

20. Yu G, Wang LG, Han Y, et al. clusterProfiler: an R package for comparing biological themes among gene clusters. *Omics* 2012;**16**(5):284-7.

21. Baron RM, Kenny DA. The moderator-mediator variable distinction in social psychological research: conceptual, strategic, and statistical considerations. *J Pers Soc Psychol* 1986;**51**(6):1173-82.

22. King HW, Orban N, Riches JC, et al. Single-cell analysis of human B cell maturation predicts how antibody class switching shapes selection dynamics. *Sci Immunol* 2021;**6**(56).

23. Takeshima Y, Iwasaki Y, Nakano M, et al. Immune cell multiomics analysis reveals contribution of oxidative phosphorylation to B-cell functions and organ damage of lupus. *Ann Rheum Dis* 2022; **81**(6):845-853.
